# Age and disc degeneration in low back pain: automated analysis enables a magnetic resonance imaging comparison of large cross-sectional cohorts of symptomatic and asymptomatic subjects

**DOI:** 10.1101/2021.11.08.21265571

**Authors:** A Jamaludin, T Kadir, A Zisserman, I McCall, FMK Williams, H Lang, E Buchanan, JP Urban, J Fairbank

**Affiliations:** Department of Engineering Science, University of Oxford, UK; Plexalis Ltd, Oxford, UK; Nuffield Department of Orthopaedics, Rheumatology and Musculoskeletal Sciences, University of Oxford, UK; Department of Twin Research and Genetic Epidemiology, King’s College London, UK; Department of Physiology, Anatomy and Genetics, University of Oxford, UK; Nuffield Orthopaedic Centre, Oxford University Hospitals NHS Trust; Emeritus, Dept of Radiology, Robert Jones and Agnes Hunt Hospital, Oswestry, UK

**Keywords:** epidemiology, herniation, Pfirrmann grading, Modic signs, stenosis

## Abstract

**Objectives:** We aimed to improve understanding of the role of imaging in diagnosis of low back pain by determining the prevalence of age-related disc degeneration in asymptomatic and symptomatic subjects. Spinal MRIs of symptomatic and asymptomatic subjects were re-annotated onto the same objective grading system and prevalence of degenerative changes compared.

**Methods:** In an exploratory cross-sectional study, we compared the prevalence of disc degeneration between two large groups of anonymised females, 30-80yrs, viz a symptomatic group with chronic back pain (724) and an asymptomatic (701) group. We used a verified automated MRI annotation system to re-annotate their spinal MRIs and report degeneration on the Pfirrmann (1-5) scale, and other degenerative changes (herniation, endplate defects, marrow signs, spinal stenosis) as binary present/absent.

**Results:** Severe degenerative changes were significantly more prevalent in discs of symptomatics than asymptomatics in the lower (L4-S1) but not the upper (L1-L3) lumbar discs in subjects <60years. We found high co-existence of several degenerative features in both populations. Degeneration was minimal in around 30% of symptomatics < 50years.

**Conclusions:** Automated MRI provides a valuable means of rapidly comparing large MRI datasets. Here, through directly comparing MRI annotations on the same objective scales it enabled us to detect significant age and spinal-level related differences in the prevalence of degenerative features between asymptomatic and symptomatic populations. By distinguishing between symptomatics whose discs have structural defects, and symptomatics with minimal degenerative changes, MRI could provide a means of clinical stratification, and provide a useful pathway to investigate possible pain sources.

**Key messages:** *What is already known about this subject?:*

- Even though intervertebral disc degeneration, and degenerative changes such as disc herniations, are strongly associated with low back pain, the importance of disc degeneration in development of low back pain is questioned because these degenerative changes are seen in both those with and those without low back pain; spinal MRIs are thus thought to be of little clinical value.

*What does this study add?:*

- The study provides the first data on age-related degeneration in those without pain and shows the significant differences in prevalence between age-related and symptom-related disc degeneration.
- The study provides definitive data showing that severe disc degeneration is directly implicated in a significant proportion of those with chronic low back pain, with the association with pain strongly dependent on age and spinal level

*How might this impact on clinical practice or future developments?:*

- The study shows that even though severe disc degeneration is strongly associated with low back pain, 30% of younger (<50yrs) chronic low back pain patients have no evident disc degeneration detected by MRI, which is important information (currently not used) for clinicians in directing treatments (and perhaps a clearer reason for the proper use of scans).
- The study provides important information for those working on mechanisms, as it enables stratification between pathways of pain arising from structural defects in the disc, and those pain pathways in discs with no such structural change.

## Introduction

Chronic low back pain is common and often difficult to treat. Its causes are mostly not well understood. Even though there is strong evidence that psycho-social factors can have a major contribution [1], the role of disc degeneration is thought to be paramount. There are over 35,000 references in PubMed on the topic ‘low back pain and intervertebral disc degeneration’ covering areas ranging from epidemiology, biological and genetics to diagnosis and treatment. Interpretation of all this information relies on adequate definitions of the disc degeneration phenotype.

Since the advent of spinal MRI permitted *in vivo* visualisation of disc degeneration, several quantitative MRI grading schemes have provided standardised definitions [2]. These have proved vital in phenotyping and analysis of genetic factors in disc degeneration [3,4], but less successful in demonstrating how disc degeneration relates to low back pain. Many asymptomatic subjects have degenerate discs [5-9] which do not necessarily predict development of future back pain [10,11]. Balagué et al., stated a widely held view: ‘Many abnormalities seen on imaging (with the possible exception of Modic signs) are equally prevalent in the asymptomatic population and merely serve as a pretext to justify overtreatment’ [12]. These views arise from prevalence data gathered from studies where gradings of disc degeneration, the degenerative features assessed and age ranges examined, vary from study to study. Systematic reviews acknowledge the difficulties of combining or comparing data while reporting presence of degenerative change [6].

Here, to provide greater insight into the prevalence of age-related disc degeneration in those with and without low back pain, **w**e report an exploratory study comparing MRIs of two symptomatic and one population group, all re-annotated onto the same rapid automated grading system (SpineNet)[13]. We compared disc degeneration between these groups in relation to age and spinal level. We examined prevalence of degenerative change in a group of female back pain patients and in a similar sized group of female asymptomatic subjects from the TwinsUK sample. We focussed on intervertebral disc degeneration, defined by the Pfirrmann grading scheme [14], and on the pathological features disc herniation, spinal stenosis, bone marrow and endplate change, all of which have been reported to be associated with symptoms[15-19].

## MATERIAL AND METHODS

### Participants and settings

All subjects were recruited with full ethical approval and consent and fully anonymised^4^ nut were not involved in study design. The study size was constrained by the groups available. As 90% of the as TwinsUK sample was female, we analysed only female subjects [20].

#### Asymptomatic Groups: (TwinsUK)

We selected 701 (69%) subjects aged 30-79 years, with no backpain during the previous 3 months (MRC Back and Neck Pain Questionnaire [4]) from the 1016 females of the TwinsUK population cohort (http://twinsuk.ac.uk) which contains both mono- and di-zygotic twin pairs. These subjects had undergone clinical examination and MRI scans (sagittal T2 sequence) with intervertebral disc degeneration graded on a 4-point scale.

#### Symptomatic Cross-sectional Group: (OSCLMRIC)

We selected 763 (39%) females, 30-79yrs from 1689 male and female patients referred to a secondary-care-centre spinal pain triage service with chronic low back pain (>3 months duration) and MR imaging.

#### Genodisc Cross-sectional Group

(https://cordis.europa.eu/project/id/201626/reporting)

Subjects were recruited in secondary care clinics in 6 different European countries. We selected the 756 (38%) females from the 2008 male and female symptomatic subjects 18-79 years, with chronic low back pain and MR imaging [9].

### Quantitative variables and MRI analysis

The sagittal T2 sequences of the MRIs from OSCLMRIC, Genodisc and TwinsUK were analysed by SpineNet, a rapid automated MRI analysis system, able to match the reliability of an experienced spinal radiologist in analysing degenerative features [13]. SpineNet takes ∼5 seconds to analyse each spine image fully, compared to around 30 minutes for manual analysis, making re-annotation of large groups feasible. It provides a report giving Pfirrmann gradings (1-5)[14] and scores degenerative features (https://zeus.robots.ox.ac.uk/spinenet2/).

For OSCLMRIC and TwinsUK, SpineNet here reported the binary characterisation of selected degenerative features, viz herniation, central canal stenosis, marrow signs (T2 weighted Sagittal T2 only), and endplate changes [13]. From this data we calculated prevalence of Pfirrmann gradings and of degenerative features in relation to spinal level and to age group by decade.

As indicated by the literature [21,19], we analysed the averages of the upper (L1/L2+ L2/L3) and lower (L4/L5+ L5/S1) lumbar intervertebral discs separately. We compared prevalence in symptomatic and asymptomatic subjects both with and without relation to age and spinal level, and calculated risks of features being symptomatic as odds ratios with 95% confidence intervals [22,23].

### Statistical methods

The prevalence of each feature by age and level in OSCLMRIC and TwinsUK (Table S5), was calculated by dividing the number of discs analyzed in each decade and spinal level with feature(s) present by the total number of discs in the same level and age group (Table S1-4). For marrow signs and endplate changes, we calculated prevalence from the average of the rostral and caudal binary scores.

Odds ratios (OR), 95% confidence intervals (CI) and P values [22,23] were used to determine the probability of an individual belonging to a group with MRI features present, being symptomatic or asymptomatic. Venn diagrams were constructed in PowerPoint®, based on the prevalence of each feature in the lower lumbar discs, with overlaps showing the prevalence of discs containing both features.

Because of the low prevalence of degenerative features in the asymptomatic group, we found that once age and disc level were considered, the study was underpowered for multivariate analysis, or for considering refinements in degradative changes such as type of herniation, degree of stenosis, or extents of endplate-defects or of marrow signs. We have used the STROBE Guidelines to structure this report.

## RESULTS

### 1. Change in mean Pfirrmann score with age and spinal level

Fig 1a,b shows the age-dependence of the rise in Pfirrmann score, averaged over the two lower (Fig 1a) and the two upper (Fig1b) lumbar discs, for two independent symptomatic groups (Genodisc, OSCLMRIC). The changes in score with age was similar for these to groups, with the lower two lumbar discs were already relatively degenerate at 30-40yrs (Fig 1a). The upper lumbar discs were only mildly degenerate at 30-40 years, but degeneration severity increased steeply with age (Fig 1b).

**Figure 1.**
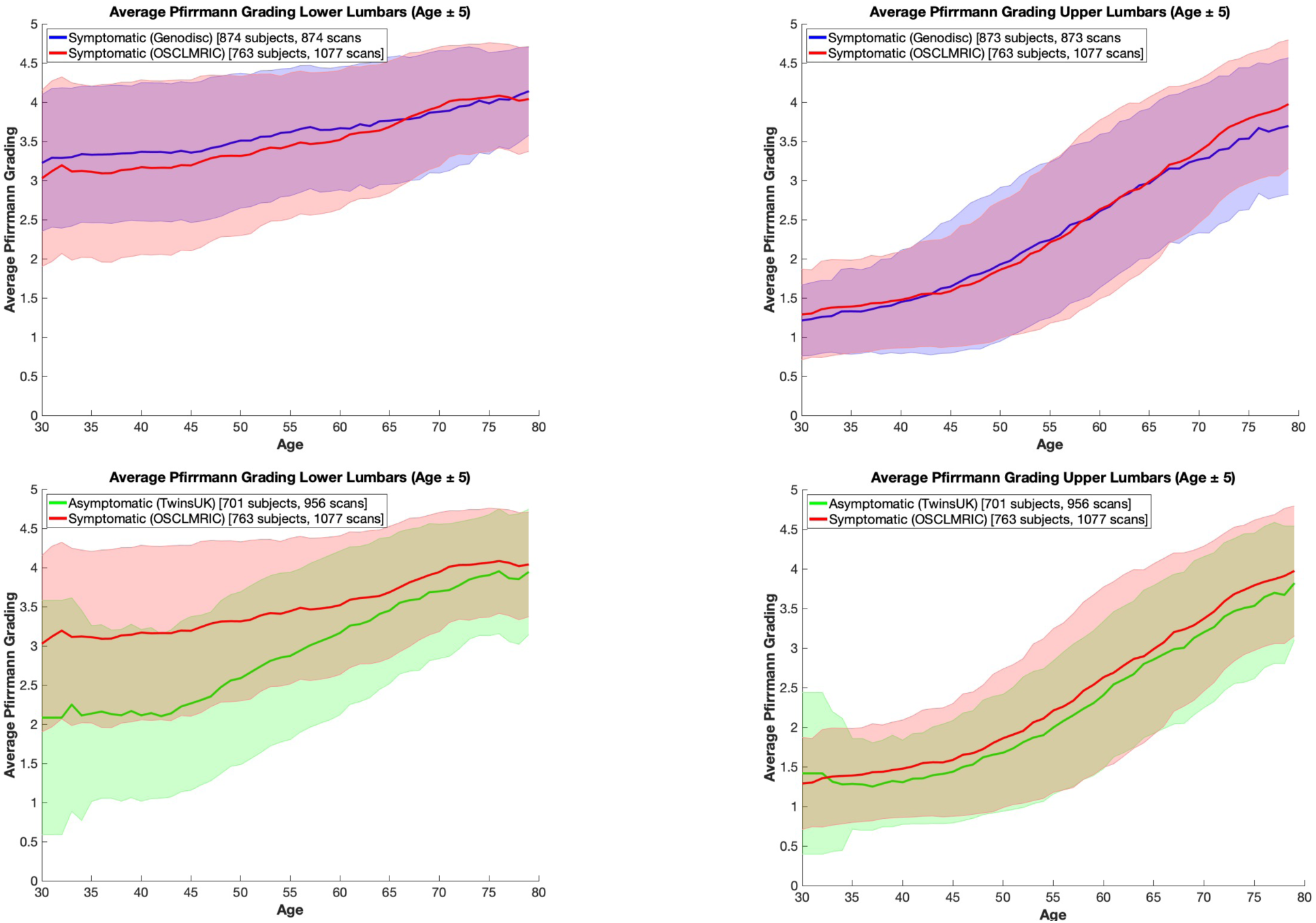
Average Pfirrmann Score versus age for lower and upper lumbar discs. A comparison between the scores in the Lower (a) and Upper (b) lumbar discs is shown for 2 patient (symptomatic) cohorts (Genodisc and OSCLMRIC). The scores of the symptomatic (OSCLMRIC female) Group are compared with an asymptomatic (Twins UK) Group in (c)for the Lower and in (d) for the Upper lumbar discs. Mean scores are shown as a solid line; one standard deviation is shown in the relevant colour and demonstrate the extent of overlap. ((Lower discs. (L4/5, L5/S1); Upper discs (L1/2, L2/3))

Figure 1c,d compares age-related Pfirrmann scores of symptomatic (OSCLMRIC) and asymptomatic (TwinsUK) subjects. In the lower lumbar discs, the average scores of symptomatics were distinctly higher than those of asymptomatics, particularly < 60yrs (Fig 1c). In the upper lumbar discs, the age-related scores were similar (Fig 1d).

### 2. Variation with age of the prevalence of high and low Pfirrmann scores for symptomatic (OSCLMRIC) and asymptomatic (TwinsUK) subjects in the lower ((L4/L5)+(L5/S1)) and upper ((L1/L2)+(L2/L3)) lumbar discs

The prevalence of severe disc degeneration (grades 4+5) in the lower lumbar discs increased with age for both groups and was 3-4 times higher in the symptomatics than the asymptomatics < 50yrs (OR >3·0; p<·0001; Fig 3b) with differences between symptomatics and asymptomatics decreasing with age (Fig 2c). In the upper lumbar spine, the prevalence was lower, and similar in the symptomatic and asymptomatic groups (Fig 2d).

**Figure 2.**
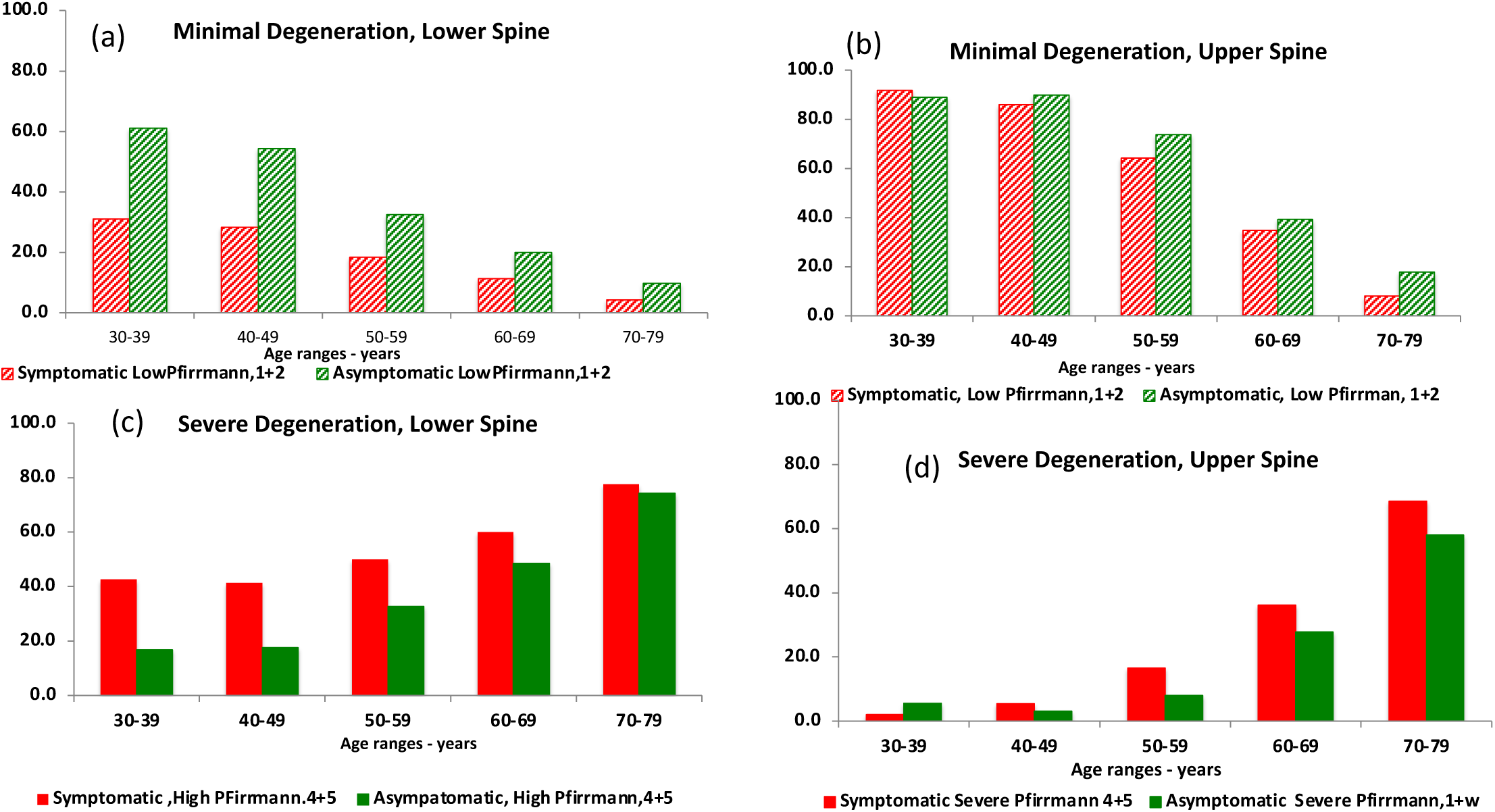
Variation with Age and Spinal Level in Prevalence (%) of Discs with High (severe) and Minimal (Low) Pfirrmann Scores between symptomatic (OSCLMRIC) and asymptomatic (TwinsUK) Subjects. This compares the prevalence of minimal (‘low; Pfirrmann 1,2’) ((a), (b)) and of severe (‘high; Pfirrmann 4,5’) ((c), (d)) degeneration between the discs of the lower ((a), (c)) and upper ((b), (d)) lumbar discs for symptomatic (OSCLMRIC) with asymptomatic subjects (TwinsUK). Lower lumbar discs (L4/5, L5/S1); Upper discs (L1/2, L2/3). (symptomatic: red; asymptomatic: green ; severe degeneration: solid; minimal degeneration: striped). Data in TableS6

**Figure 3.**
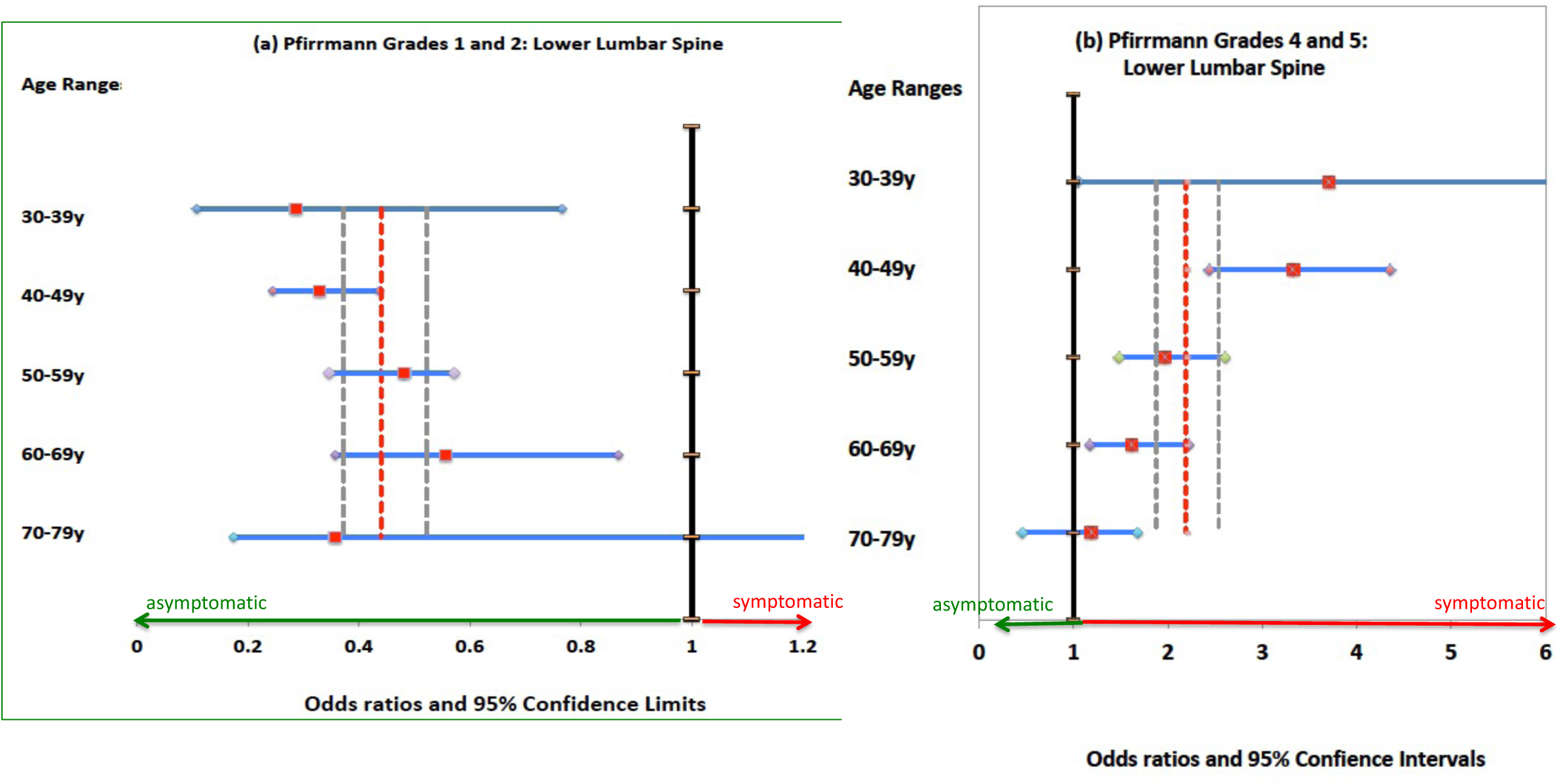
Odds Ratios and 95% Confidence Intervals for disc degeneration scores by age, both pooled and by decade; symptomatics (OSCLMRIC) vs. asymptomatics (TwinsUK) for low (1 or 2) (a) and high (4 or 5) (b) Pfirrmann scores in the low lumbar spine (L4/5, L5/S1). Vertical broken line is OR for pooled data (Green broken vertical lines are 95% CI.) Data in Table 5b

The prevalence of minimally degenerate discs in the lower lumbar spine was significantly greater in asymptomatics over the whole age range (Fig 2a) but fell steeply with age for both groups with no significant difference between groups by 70yrs (Fig 3a).

The prevalence of severely degenerate discs over the whole lumbar spine, disregarding age and spinal level, was 37% for the symptomatic and 23% for the asymptomatics. Though differences between symptomatics and asymptomatics were significant (OR 1·94: 95% CI:1·48-1.55, p<·0001), pooling these data lost information on the influence of age and spinal level.

### 3. Prevalence with age and disc level of degenerative features in symptomatic and asymptomatic subjects

The prevalence of disc herniation (Fig 4a), central canal stenosis (Fig 4b), marrow change (Fig 4c) and endplate defect (Fig 4d) varied with age and disc level. For all features, prevalence was markedly greater in symptomatic than in asymptomatic subjects, particularly at younger age, and, apart from endplate defect, greater in the lower lumbar spine. For instance, in the lower lumbar spine at 40-49 years, around 30% of discs were herniated in symptomatics compared to 9% of asymptomatic discs (p <·0001, Fig 4a,Table 6b). Over the same age range, marrow change was present in 32% of symptomatic compared to 11% of asymptomatic discs (p<·00001, Fig 4c; Table S6b); for endplate defects, prevalence was lower being 8% for symptomatics and 3% in asymptomatics (p<·00001, Fig 4d, Table S6b)

**Figure 4.**
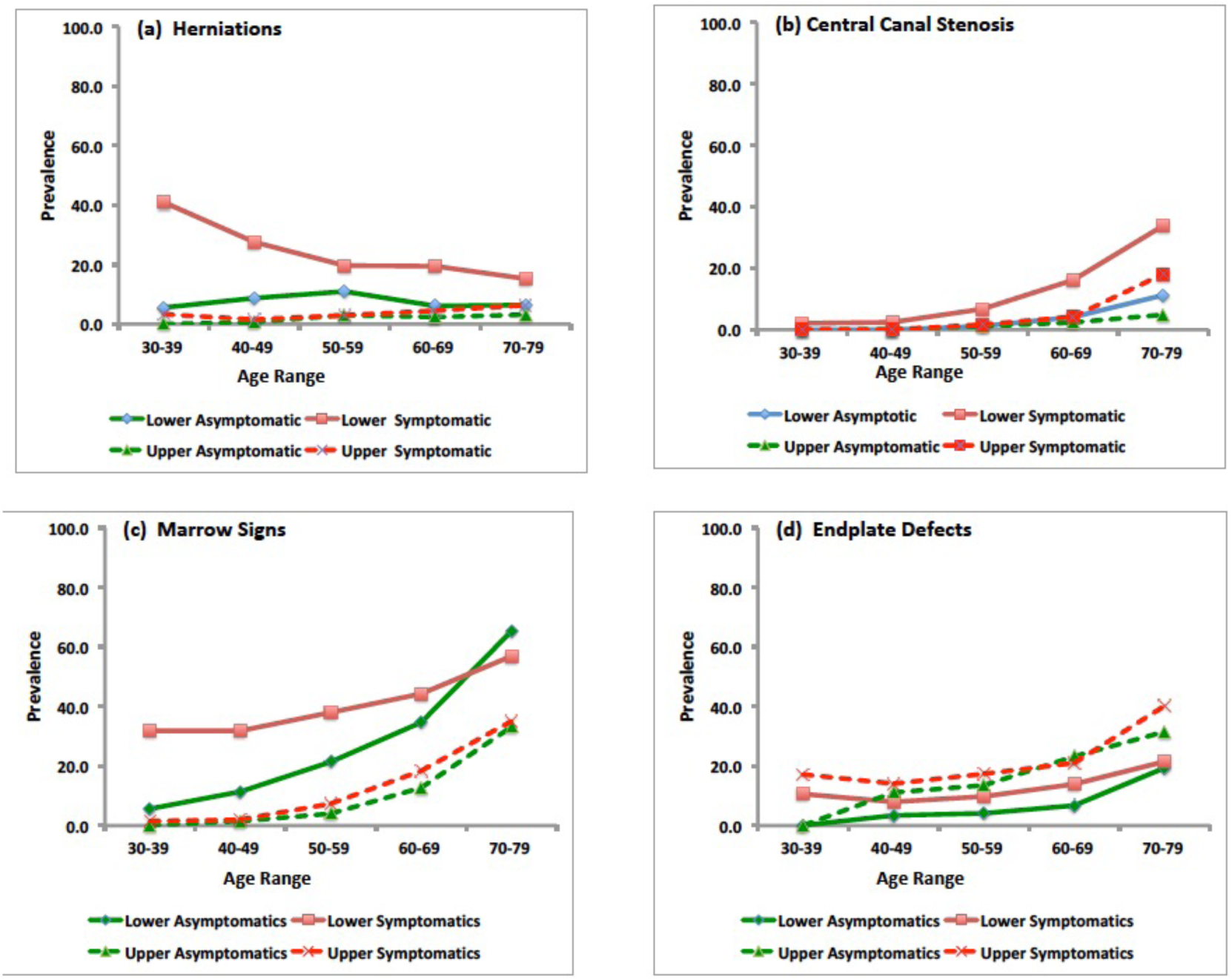
Prevalence of degenerative features in relation to age pooled and by decade; symptomatic vs asymptomatic cohorts. The figures show the prevalences of (a) herniations (b) central canal stenosis (c) marrow signs (d) endplate defects in the lower and upper lumber discs in relation to ages ranges, shown by decade. (Symptomatic: red lines, asymptomatic: green lines: lower spine solid lines, upper spine, broken lines). Data shown in Table S6.

The prevalence of herniation, marrow changes and central canal stenosis in the upper lumbar discs was very low, <10%, and was similar in both groups (Tables S5). For endplate defects however, prevalence was greater in the upper spine, similar in both groups and increased with age (Fig. 4d; Table S5)

The lower lumbar ORs were strongly age dependent (Fig 5a-d;Table S6b). For herniation in the lower lumbar discs for instance, the OR varied from 11·8 (95% CI 1·5-90·0; p=0·02) to 2·1 (95% CI;1·4-3·1; p<·00001) and then rose to 3·7(95%CI 2·2-6·2; p<·00001) in the 4^th^, 6^th^ and 7th decades respectively (Fig 5, Table S6b); confidence intervals were wide because of the small number of herniations in asymptomatic subjects. Overall, the prevalence of degenerative features was significant (Table S6a), but the effects of age and disc level were lost.

**Figure 5.**
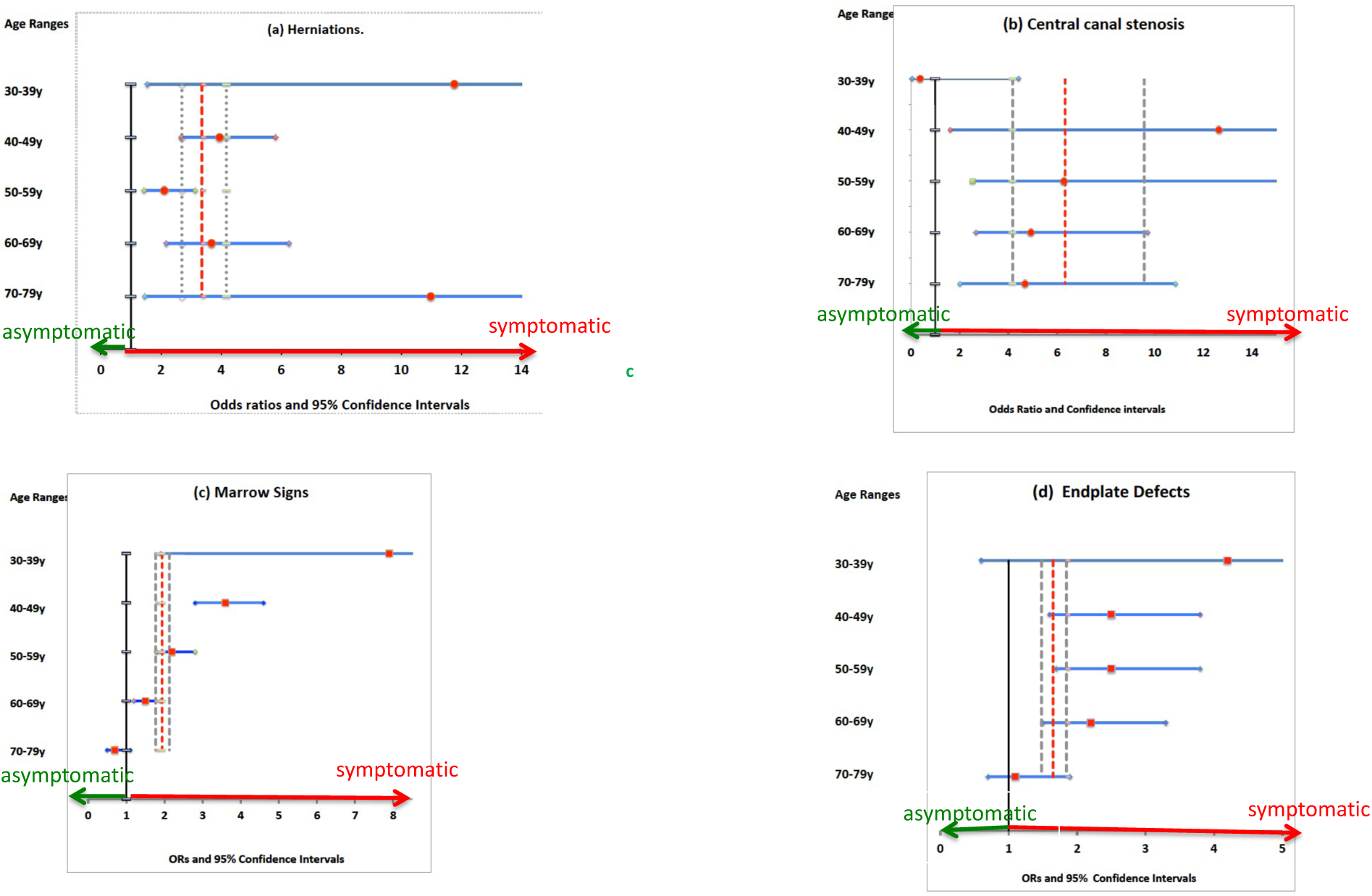
Odds Ratios and Confidence Intervals for prevalence of degenerative features in relation to age; symptomatic vs asymptomatic groups. The Odds ratios for prevalence of degenerative features in symptomatic vs. asymptomatic subjects are shown by age, with (years) both pooled and by decade. The red squares show the odds ratio and the blue line show 95% confidence intervals for the degenerative features in the low lumbar discs (L4/5, L5/S1) per decade (30-80 years) for(a) herniations (b) central canal stenosis (c) marrow changes (d) endplate changes in the symptomatic versus asymptomatic subjects. The vertical red broken lines show the pooled odds ratios and the grey dotted lines the overall confidence intervals, including all ages and spinal levels for (a) herniations (b) central canal stenosis (c) marrow signs (d) endplate defects in the symptomatic versus asymptomatic cohorts. Herniations predominate in the young, whilst degenerative stenosis predominates in the elderly. Developmental stenosis is detected in the young symptomatics (prevalence ∼2%), but not the asymptomatics. Data shown in Table S5b

### 4. Most degenerative features are in discs with high Pfirrmann scores

The majority of degenerative features, in both symptomatics and asymptomatics, were found in discs with Pfirrmann grades 4 or 5 (Fig 6 a-d) whatever the overall prevalence of the feature. More than 90% of marrow changes were seen in Pfirrmann 4 or 5 discs for both symptomatic and asymptomatic subjects at all ages. The low (3%) prevalence of spinal stenosis in symptomatics < 50 years, had no apparent association with disc degeneration, possibly reflecting ‘developmental’ rather than ‘degenerative’ stenosis [24] (Fig 4b. Table S5).

**Figure 6.**
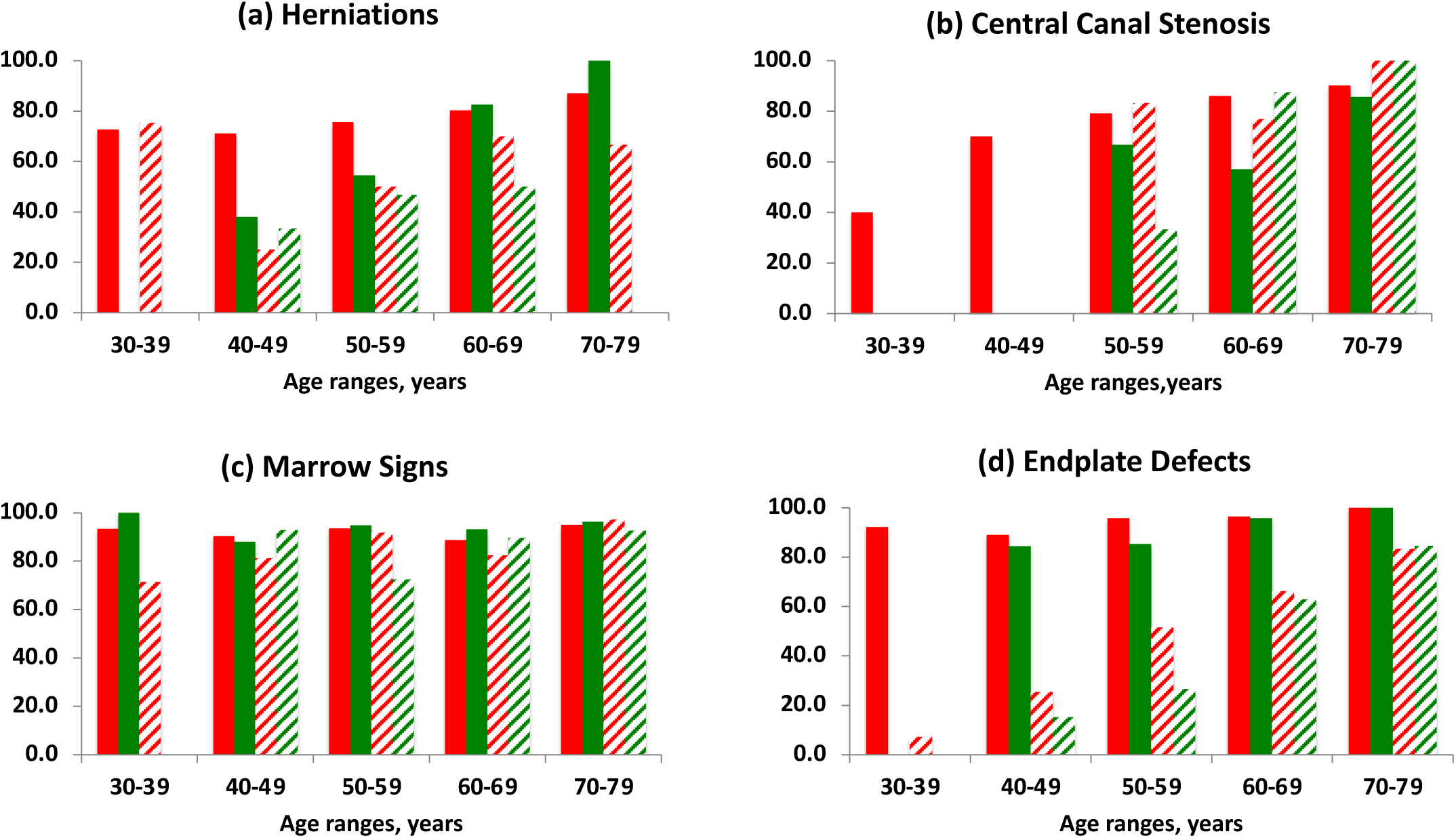
Age-related Percentage of lower lumbar discs from the symptomatic and asymptomatic cohorts which have specific degenerative features, and which are Pfirrmann Grades 4 or 5. The percent of all lower lumbar discs which have the specific degenerative features (a) herniations, (b) central canal stenosis (c) endplate defects (d) marrow signs and also have severe disc degeneration (Pfirrmann 4+5) are shown in relation to age for the symptomatic (red) and asymptomatic (green) groups. Data calculated from Tables S1-4.

### 5 Co-existence of degenerative features

As shown by a Venn diagram (Fig 7), for 40-49y subjects, around 14% of symptomatics and 12·3% of asymptomatics had only a single degenerative feature; marrow signs alone were seen in only 0.5% of symptomatic discs. Similar co-existence of degenerative features was seen at all ages (Tables S1-S5). At 30-60 years, around 30% of the symptomatic group had no/mild degeneration (Pfirrmann 1 or 2), and no identified degenerative features (Table 1).

**Figure 7.**
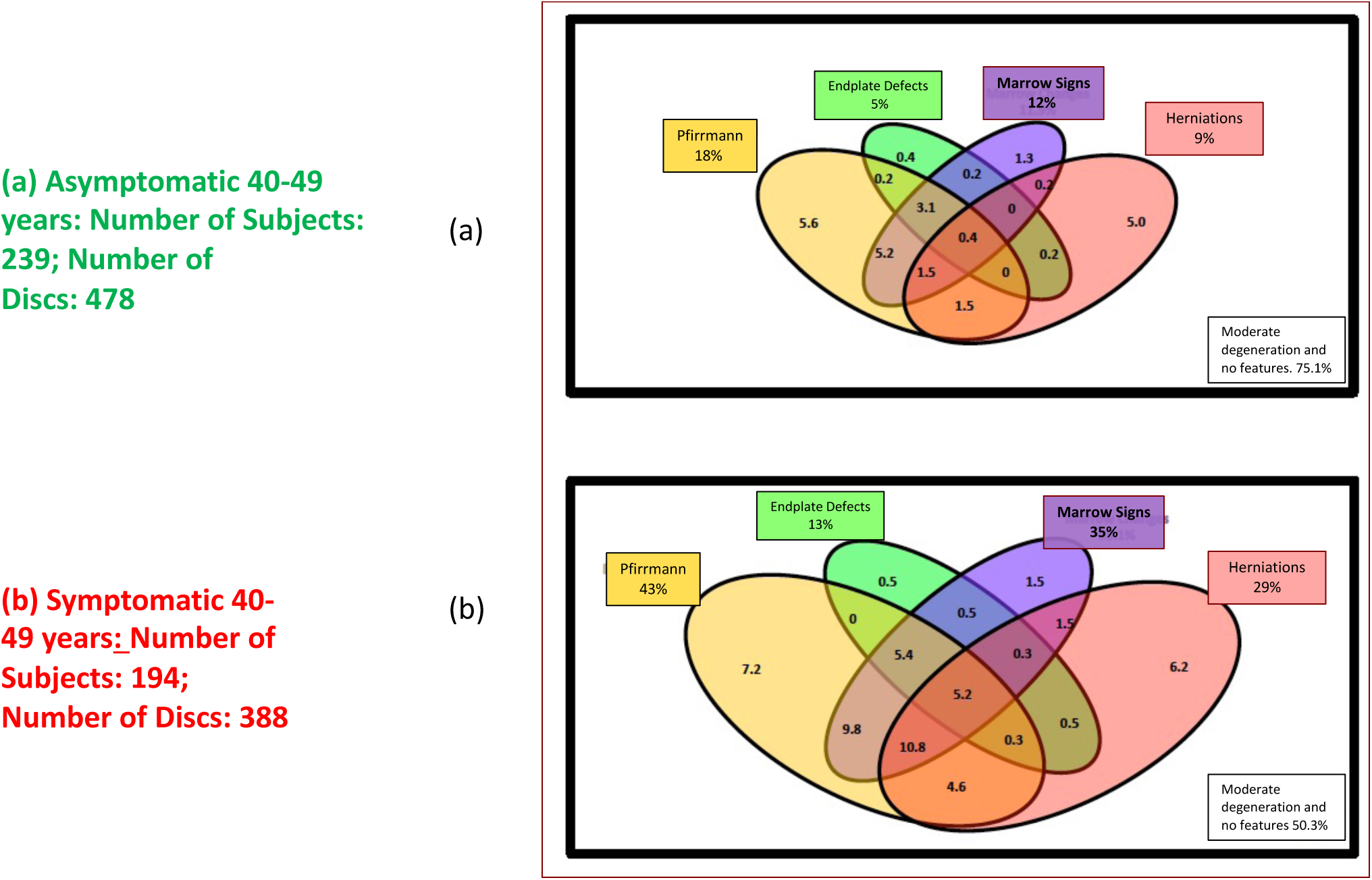
Venn diagrams, showing the prevalence (%) and overlap of degenerative features for symptomatic and asymptomatic Lower Lumbar discs, 40-49yrs. The prevalence of degenerative features is shown here for (a) symptomatic discs (388) and (b) asymptomatic discs(478) and gives the prevalence of severe disc degeneration (Pfirrmann 4+5), herniations, endplate defects and marrow changes and also the percentage of discs which have no degenerative features and are only mildly degenerate. Each degenerative feature is colour-coded. The total prevalence of each feature is shown outside the diagram and the percent overlap between features is shown inside the Venn diagrams. The prevalence of central canal stenosis is very low in this age group (Fig. 4) so is not shown and may reflect developmental rather than degenerative stenosis. Data taken from Tables S1-4, Table S6.

**Table 1;.**
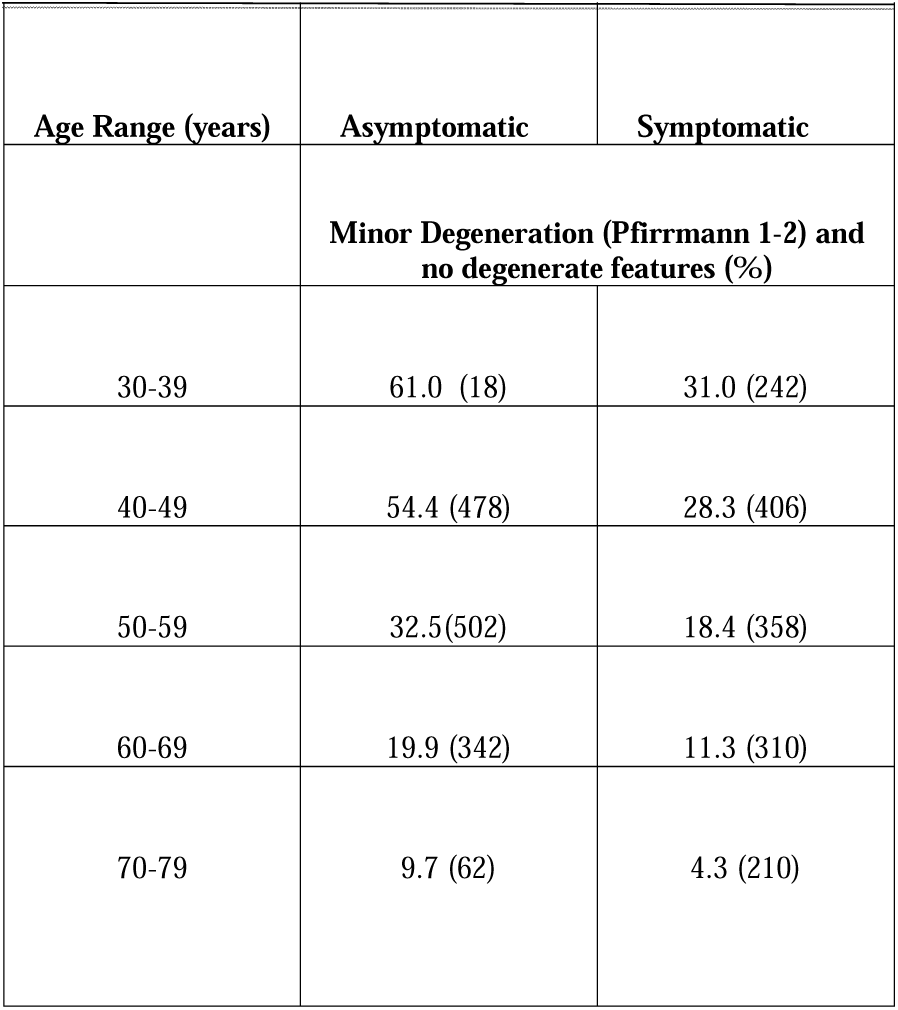
Percentage of discs with mild degeneration and no identified degenerative features in relation to age;. lower lumbar ((L4/L5) and (L5/S1)) discs which have no degenerative features identified here (viz. herniations,
central canal stenosis, marrow signs, endplate defects) and only mild degeneration (Pfirrmann 1+2).

## DISCUSSION

Here we show that an automated MRI analysis system (SpineNet), enabled images of large groups to be re-annotated rapidly onto the same grading system independent of their original annotations, and hence compared in relation to age- and spinal level-related degenerative changes.

By comparing annotated images from lumbar spines of asymptomatic subjects (TwinsUK) with those from symptomatic subjects with chronic low back pain (OSCLMRIC), we found, in contrast to published view[12], the prevalence of severe disc degeneration (Fig 2c) and of degenerative features such as herniations and marrow signs were not similar but were markedly greater in the lower lumbar discs of symptomatics than in those of asymptomatics (Fig4), with significant age-dependent odds-ratios (Fig 3, Fig 5). In the upper lumbar spine however, there was no significant difference between asymptomatics and symptomatics for any of the features examined, and the prevalence was far lower than in the lower lumbar spine <70yrs (Fig 2d). If age and spinal level were not taken into account, the odds ratios were still significant (Table S6a), but information on the effects of age and spinal level on their magnitude was lost.

We found several degenerative changes tended to co-exist in discs of both symptomatic and asymptomatic subjects (Fig 7a,b). Moreover, more than 90% of discs with marrow signs, and a high proportion of disc with herniations or endplate defects were seen in severely degenerate discs in both symptomatic and asymptomatic subjects over all age ranges (Figs 6,7). While marrow signs and endplate changes have each been associated singly with symptoms [25,16,17], in view of their high co-existence with severe degeneration, it is questionable if these defects alone are a liikely source of pain. Strong associations with pain have been found when several co-existing degenerative features are considered together [15,26].

Why degenerative changes appear to be strongly related to pain in some subjects [27], and why others with apparently similar degenerative changes remain pain free is still unclear. Some studies, mostly small, have found that the type of herniation [28], endplate defect [16], Modic change [17] or degree of stenotic change [18] can distinguish images of symptomatic from those of asymptomatic discs. Much larger numbers of subjects than those examined here will be needed to test these findings if effects of age and disc level are to be considered. Unbiased analyses of degeneration, or MRI sequences other than T2, might also be better able to discriminate between painful and non-painful discs [29]. However even advances in MRI imaging protocols [30] may not be able to differentiate between some asymptomatic and symptomatic subjects. Load-induced effects on other spinal structures [31,32,33] cannot be detected by conventional supine MRIs but axially loaded MRIs appear able to differentiate between those with and without pain in regard, for instance, to images of the spinal canal dimensions or endplate features [34,35]. Also, current conventional MRIs are unable to detect augmented pain processing or psycho-social factors, also associated with low back pain [1,36,37].

Around 30% of subjects 30-50yrs, had chronic backpain but no apparent degenerative changes (Table 1). As pain could be induced by different pathways in those with structural changes to their discs visible by imaging and those without, spinal MRIs able to detect degree of degenerative changes, have potential clinical and research benefit.

## Limitations

Our asymptomatic group, TwinsUK, only had subjects 30-79 years. The OSCLMRIC group were chronic back-pain patients, referred to secondary care mainly through failure to respond to interventions in primary care, effectively excluding many subjects who have a natural resolution of their symptoms and those referred to pain clinics. These patients may be distinct from the totality of patients seen in primary care, such as those discussed in the Lancet series [38], or those in a population cohort which found no differences in degenerative changes between those with and without low back pain [8]. A similar pattern of degenerative change (Fig1a) was seen in OSCLMRIC subjects and those of Genodisc, another secondary care group. Patients recruited in other settings could differ in definitions of pain symptoms, gender and clinical subtypes.

The prevalence of degenerative changes reported here is specific to our study. Reports of degeneration vary from study to study. In a rural farming population in Japan, the prevalence of severe disc degeneration (Pfirrmann 4 or 5) in the lower lumbar spine of asymptomatic women was ∼70% at 50-69 years, and ∼80% at 60-69years[39], compared to ∼33% and ∼45% respectively to the females in TwinsUK (Fig 2c). The prevalence of degenerative features, such as herniations in asymptomatics, also varies, from around 63% at 20-50 years [40] to 22% at 40-49 years [5] compared to <10% over the same age ranges in our study (Fig 4a). Differences in annotation could also affect reported prevalence [41]. Consistent annotation of large groups is critical to progressing our understanding of these complex relationships.

A limitation likewise lies in the imaging protocols used here. We saw a similarity in Pfirrmann scores of two different patient groups annotated by SpineNet (Fig 1) despite the differences in MR machines used [13], but as Twins UK had only Sagittal T2 examinations, here we reported only these sequences. Other imaging sequences and refinement of annotations of degenerate features may provide information which is better able distinguish painful from non-painful discs [42] but will require larger study groups if influences of age and disc level are considered.

Our analysis of degeneration uses 1) an average of the 1-5 categoric Pfirrmann scale (Fig 1), which conceals important information, and 2) an analysis of grades 1 or 2 versus 4 or 5 which identifies populations with either disc ‘normal’ or disc ‘very degenerate’. It omits changes in grade 3 discs, around 25% of the total discs in both groups at some ages (Tables S5). However, most of the other degenerative features, predominantly seen in ‘very abnormal’ discs (Fig 6) were not omitted. Larger data sets could allow a sensitivity analysis of these issues.

## Conclusions

Automated annotation can overcome the considerable challenges facing experts in annotating complex imaging in large groups [43]. Moreover, by rapidly re-annotating large groups, previously annotated by different MRI analysis systems on the same objective system, it enables data from large pre-existing groups to be compared or combined. Collection of new groups is expensive and time consuming. Automated analysis provides a way in which epidemiological analysis could be advanced by combining and comparing data from existing groups with MRIs and information on back pain.

Here we demonstrated that an automated analysis system (SpineNet) enabled rapid re-annotation on the same grading system, of two chronic low back pain patient groups (Genodisc and OSCLMRIC) and a population sample drawn from the TwinsUK cohort previously annotated by different coding methods. We found age and spinal level influenced degenerative changes strongly. Discs of symptomatics in the lower lumbar spine of chronic low back pain patients<70years, were significantly more degenerate than age-matched discs of those with no pain, in agreement with a systematic review [44]. In the upper lumbar spine of those <70 years, degenerative changes were mainly far less prevalent and were similar in symptomatics and asymptomatics. Hence reporting MRI features of spinal pathology in relation to symptoms, without taking age or spinal level into account, can be very misleading. We suggest information on age and disc level should be incorporated in clinical reporting standards to maximise the value of current NHS usage of MRI scans in this population.

In our study, 30% of patients aged 30-50 years with back pain (compared with 60% without back pain) had no degenerative features detectable on MRI. We suggest that spinal MRIs, by distinguishing between spines with and without structural degenerative changes, could provide a means of stratifying patients for further research into causes and treatments of low back pain.

## Data Availability

All data produced in the present work are contained in the manuscript

## The Patient and Public Involvement statement

- At what stage in the research process were patients/the public first involved in the research and how? Automated annotation has been presented through BackCare patients’ charity and in a public question and answer session at the Back Pain Show, Birmingham May 2017 and subsequent interactive podcasts. The symptomatic group and asymptomatic cohort have had PPI input in their clinical and research settings. The Oxford spine clinic is a serial recruiting setting for clinical studies with regular interactions with large numbers of patients concerning back pain research (EB).
- How were the research question(s) and outcome measures developed and informed by their priorities, experience, and preferences? MRI is considered essential by many patients and current guidance discourages the use of imaging for the investigation of back pain (EB)
- How were patients/the public involved in the design of this studyã Not directly, but epidemiology issues were raised in public discussions
- How were they involved in the recruitment to and conduct of the studyã These subjects were recruited anonymously based on reporting back pain and having suitable imaging. TwinsUK has a full explanation of its history and recruitment on its website: https://twinsuk.ac.uk/ It is stated in the Methods that subjects were not involved in the design of this study.

## Funding sources

SpineNet: EPSRC Programme Grant Seebibyte (EP/M013774/1)

Genodisc: EC FP7 project GENODISC (HEALTH-F2-2008-201626)

TwinsUK: Funded by the Wellcome Trust; European Community’s Seventh Framework Programme (FP7/2007– 2013)

OSCLMRIC: Funded by Back-to-Back Charity (1079089)

None of these funding sources have been involved in the writing of the manuscript or the decision to submit it for publication.

## Figure and Table Legends

**Table 1.**

**Percentage of Discs with No Identified Degenerative Features in relation to Age; Lower lumbar asymptomatic and symptomatic discs.** The percentage of asymptomatic and symptomatic lower lumbar ((L4/L5) and (L5/S1)) discs which have no degenerative features identified here (viz. herniations, central canal stenosis, marrow signs, endplate defects) and only mild degeneration (Pfirrmann 1+2) or moderate degeneration (Pfirrmann 1+2+3) with disc numbers analysed given in parentheses.

## Supplementary Data

**Tables S1-S4**.

**The information over each age group on the number of discs examined**.

Information is given for both the lower lumbar spine ((L4/5) and L5/S1)) and the upper lumbar spine ((L1/L2) and (L2/L3) on the number of discs of each Pfirrmann grade, and the number of discs with each feature examined for both the aymptomatic (a) and the asymptomatic (b) groups. S1 gives information on numbers of discs with herniations, S2 with marrow changes, S3 with endplate defects, S4 with central canal stenosis.

**Table S1a.**

**The number of discs in the symptomatic group found with herniations**. Results are shown in relation to spinal levels (lower lumbar or upper lumbar), age range and Pfirrmann gradings.

**Table S1b.**

**The number of discs in the asymptomatic group found with herniations**. Results are shown in relation to spinal levels (lower lumbar or upper lumbar), age range and Pfirrmann gradings.

**Table S2a.**

**The number of end plates in the symptomatic group found with marrow changes**. Results are shown in relation to spinal levels (lower lumbar or upper lumbar), age range and Pfirrmann gradings.

**Table S2b.**

**The number of end plates in the asymptomatic group found with marrow changes**. Results are shown in relation to spinal levels (lower lumbar or upper lumbar), age range and Pfirrmann gradings.

**Table S3a.**

**The number of end plates in the symptomatic group found with endplate defects**. Results are shown in relation to spinal levels (lower lumbar or upper lumbar), age range and Pfirrmann gradings.

**Table S3b.**

**The number of end plates in the asymptomatic group found with endplate defects**. Results are shown in relation to spinal levels (lower lumbar or upper lumbar), age range and Pfirrmann gradings.

**Table S4a.**

**The number of discs in the symptomatic group found with central canal stenosis**. Results are shown in relation to spinal levels (lower lumbar or upper lumbar), age range and Pfirrmann gradings.

**Table S4b.**

**The number of discs in the asymptomatic group found with central canal stenosis**. Results are shown in relation to spinal levels (lower lumbar or upper lumbar), age range and Pfirrmann gradings.

**Table S5a.**

**Odds ratios (OR) (symptomatic/asymptomatic), confidence intervals (CI) and probability estimates (P), values for each feature, without considering age or spinal level.**

**Table S5b.**

**Odds ratios (OR) (symptomatic/asymptomatic), P confidence intervals (CI) and probability estimates (P) values for each feature, taking into account age, shown here for the lower lumbar spine** (Discs of the lower lumbar spine (L4/L5) and (L5/S1)).

**Table S6. Prevalences of degenerative features in regard to age and spinal level for symptomatic (a) and asymptomatic (b) subjects.** Prevalences are shown for Pfirrmann (1+2), Pfirrmann 3, Pfirrmann 4+5, herniations, central canal stenosis, marrow signs, and endplate defects in relation to age for the lower (L4/L5,L5/S1) and upper (L1/L2, L2/L3) lumbar discs.

**Table S1a.** The number of discs in the symptomatic group found with herniations. Results are shown in relation to spinal levels (lower lumbar or upper lumbar), age range and Pfirrmann gradings.

| Symptomatic (OSCLMRIC) - Pfirrmann and herniations |  |  |  |  |  |  |  |  |  |  |  |  |
| --- | --- | --- | --- | --- | --- | --- | --- | --- | --- | --- | --- | --- |
| MRI Imaging features | Lower Lumbar (L5/S1, L4/L5) |  |  |  |  |  | Upper Lumbar (L1/L2, L2/L3) |  |  |  |  |  |
| Age ranges | 30-39 | 40-49 | 50-59 | 60-69 | 70-79 | Total: | 30-39 | 40-49 | 50-59 | 60-69 | 70-79 | Total: |
| Total number of subjects | 121 | 203 | 179 | 155 | 105 | 763 | 121 | 203 | 179 | 155 | 105 | 763 |
| Total number of discs | 242 | 406 | 358 | 310 | 210 | 1526 | 242 | 406 | 358 | 310 | 210 | 1526 |
| Number of discs with Pfirrmann-1,2 | 75 | 115 | 66 | 35 | 9 | 300 | 222 | 349 | 230 | 108 | 17 | 926 |
| Number of discs with Pfirrmann-3 | 64 | 123 | 113 | 89 | 38 | 427 | 15 | 35 | 69 | 90 | 49 | 258 |
| Number of discs with Pfirrmann-4,5 | 103 | 168 | 179 | 186 | 163 | 799 | 5 | 22 | 59 | 112 | 144 | 342 |
| Number of Discs with Herniations | 99 | 114 | 74 | 61 | 31 | 379 | 8 | 6 | 10 | 15 | 12 | 51 |
| Number of discs Pfirrmann-1,2 with herniations | 3 | 1 | 0 | 2 | 0 | 6 | 2 | 2 | 0 | 0 | 0 | 4 |
| Number of discs Pfirrmann-3 with herniations | 24 | 32 | 18 | 10 | 4 | 88 | 4 | 1 | 3 | 5 | 2 | 15 |
| Number of discs Pfirrmann-4,5 with herniations | 72 | 81 | 56 | 49 | 27 | 285 | 2 | 3 | 7 | 10 | 10 | 32 |

**Table S1b.**
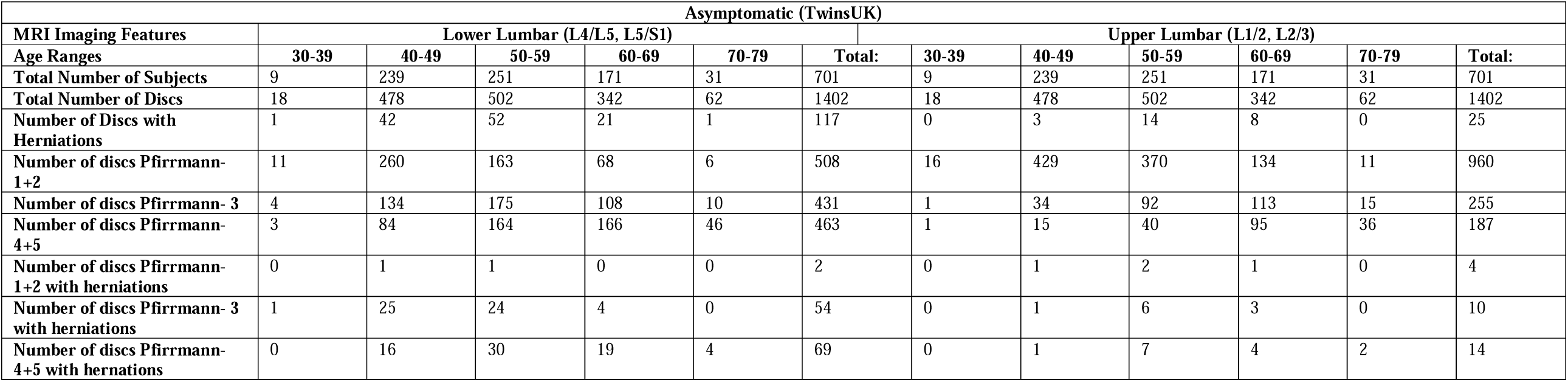
The number of discs in the asymptomatic group found with herniations. Results are shown in relation to spinal levels (lower lumbar or upper lumbar), age range and Pfirrmann gradings.

**Table S2a.** The number of end plates in the symptomatic group found with marrow changes. Results are shown in relation to spinal levels (lower lumbar or upper lumbar), age range and Pfirrmann gradings.

| Symptomatics (OSCLMRIC) |  |  |  |  |  |  |  |  |  |  |  |  |
| --- | --- | --- | --- | --- | --- | --- | --- | --- | --- | --- | --- | --- |
| MRI Imaging features | Lower Lumbar (L5/S1, L4/L5) |  |  |  |  |  | Upper Lumbar (L1/L2, L2/L3) |  |  |  |  |  |
| Age ranges | 30-39 | 40-49 | 50-59 | 60-69 | 70-79 | Total: | 30-39 | 40-49 | 50-59 | 60-69 | 70-79 | Total: |
| Total number of subjects | 121 | 203 | 179 | 155 | 105 | 763 | 121 | 203 | 179 | 155 | 105 | 763 |
| Total number of endplates | 484 | 812 | 716 | 620 | 420 | 3052 | 484 | 812 | 716 | 620 | 420 | 3052 |
| Number of endplates adjacent to discs with Pfirrmann-1,2 | 150 | 230 | 132 | 70 | 18 | 600 | 444 | 698 | 460 | 216 | 34 | 1852 |
| Number of endplates adjacent to discs with Pfirrmann-3 | 128 | 246 | 226 | 178 | 76 | 854 | 30 | 70 | 138 | 180 | 98 | 516 |
| Number of endplates adjacent to discs with Pfirrmann-4,5 | 206 | 336 | 358 | 372 | 326 | 1598 | 10 | 44 | 118 | 224 | 288 | 684 |
| Number of Endplates with Marrow Signs | 154 | 258 | 263 | 274 | 239 | 1188 | 7 | 16 | 49 | 114 | 147 | 333 |
| Number of Endplates with marrow signs adjacent to discs with Pfirrmann-1,2 | 0 | 2 | 0 | 1 | 1 | 4 | 0 | 1 | 1 | 4 | 0 | 6 |
| Number of Endplates with marrow signs adjacent to discs with Pfirrmann-3 | 10 | 23 | 17 | 30 | 11 | 91 | 2 | 2 | 3 | 16 | 4 | 27 |
| Number of Endplates with marrow signs adjacent to discs with Pfirrmann-4,5 | 144 | 233 | 246 | 243 | 227 | 1093 | 5 | 13 | 45 | 94 | 143 | 300 |

**Table S2b.** The number of end plates in the asymptomatic group found with marrow changes. Results are shown in relation to spinal levels (lower lumbar or upper lumbar), age range and Pfirrmann gradings.

| Asymptomatic (TwinsUK) |  |  |  |  |  |  |  |  |  |  |  |  |
| --- | --- | --- | --- | --- | --- | --- | --- | --- | --- | --- | --- | --- |
| MRI Imaging Features | Lower Lumbar (L4/L5, L5/S1) |  |  |  |  |  | Upper Lumbar (L1/2, L2/3) |  |  |  |  |  |
| Age Ranges | 30-39 | 40-49 | 50-59 | 60-69 | 70-79 | Total: | 30-39 | 40-49 | 50-59 | 60-69 | 70-79 | Total: |
| Total Number of Subjects | 9 | 239 | 251 | 171 | 31 | 701 | 9 | 239 | 251 | 171 | 31 | 701 |
| Total Number of Endplates | 36 | 956 | 1004 | 684 | 124 | 2804 | 36 | 956 | 1004 | 684 | 124 | 2804 |
| Number of Endplates with Marrow signs | 2 | 109 | 215 | 237 | 81 | 644 | 0 | 14 | 40 | 87 | 41 | 182 |
| Number of Discs with Pfirrmann-1+2 | 11 | 260 | 163 | 68 | 6 | 508 | 16 | 429 | 370 | 134 | 11 | 960 |
| Number of Discs with Pfirrmann-3 | 4 | 134 | 175 | 108 | 10 | 431 | 1 | 34 | 92 | 113 | 15 | 255 |
| Number of Discs with Pfirrmann-4+5 | 3 | 84 | 164 | 166 | 46 | 463 | 1 | 15 | 40 | 95 | 36 | 187 |
| Number of endplates adjacent to disc Pfirrmann- 1+2 and Marrow signs | 0 | 0 | 0 | 2 | 0 | 2 | 0 | 0 | 1 | 1 | 0 | 2 |
| Number of endplates adjacent to disc Pfirrmann-3 with Marrow signs | 0 | 13 | 11 | 14 | 3 | 41 | 0 | 1 | 10 | 8 | 3 | 22 |
| Number of endplates adjacent to disc Pfirrmann- 4+5 with Marrow signs | 2 | 96 | 204 | 221 | 78 | 601 | 0 | 13 | 29 | 78 | 38 | 158 |

**Table S3a.** The number of end plates in the symptomatic group found with endplate defects. Results are shown in relation to spinal levels (lower lumbar or upper lumbar), age range and Pfirrmann gradings.

| Symptomatic (OSCLMRIC) |  |  |  |  |  |  |  |  |  |  |  |  |
| --- | --- | --- | --- | --- | --- | --- | --- | --- | --- | --- | --- | --- |
| MRI Imaging features | Lower Lumbar (L5/S1, L4/L5) |  |  |  |  |  | Upper Lumbar (L1/L2, L2/L3) |  |  |  |  |  |
| Age ranges | 30-39 | 40-49 | 50-59 | 60-69 | 70-79 | Total: | 30-39 | 40-49 | 50-59 | 60-69 | 70-79 | Total: |
| Total number of subjects | 121 | 203 | 179 | 155 | 105 | 763 | 121 | 203 | 179 | 155 | 105 | 763 |
| Total number of endplates | 484 | 812 | 716 | 620 | 420 | 3052 | 484 | 812 | 716 | 620 | 420 | 3052 |
| Number of endplates adjacent to discs with Pfirrmann-1,2 | 150 | 230 | 112 | 70 | 18 | 580 | 444 | 698 | 460 | 216 | 34 | 1852 |
| Number of endplates adjacent to discs with Pfirrmann-3 | 128 | 246 | 226 | 178 | 76 | 854 | 30 | 70 | 138 | 180 | 98 | 516 |
| Number of endplates adjacent to discs with Pfirrmann-4,5 | 206 | 336 | 358 | 372 | 326 | 1598 | 10 | 44 | 118 | 224 | 288 | 684 |
| Number of Endplates with Defects | 51 | 64 | 70 | 86 | 90 | 361 | 83 | 114 | 124 | 130 | 169 | 620 |
| Number of Endplates with Defects adjacent to discs with Pfirrmann-1,2 | 2 | 1 | 1 | 0 | 0 | 4 | 60 | 58 | 30 | 9 | 7 | 164 |
| Number of Endplates with Defects adjacent to discs with Pfirrmann-3 | 2 | 6 | 2 | 3 | 0 | 13 | 17 | 27 | 30 | 35 | 21 | 130 |
| Number of Endplates with Defects adjacent to discs with Pfirrmann-4,5 | 47 | 57 | 67 | 83 | 90 | 344 | 6 | 29 | 64 | 86 | 141 | 326 |

**Table S3b.** The number of end plates in the asymptomatic group found with endplate defects. Results are shown in relation to spinal levels (lower lumbar or upper lumbar), age range and Pfirrmann gradings.

| Asymptomatic (TwinsUK) |  |  |  |  |  |  |  |  |  |  |  |  |
| --- | --- | --- | --- | --- | --- | --- | --- | --- | --- | --- | --- | --- |
| MRI Imaging Features | Lower Lumbar (L4/L5, L5/S1) |  |  |  |  |  | Upper Lumbar (L1/2, L2/3) |  |  |  |  |  |
| Age Ranges | 30-39 | 40-49 | 50-59 | 60-69 | 70-79 | Total: | 30-39 | 40-49 | 50-59 | 60-69 | 70-79 | Total: |
| Total Number of Subjects | 9 | 239 | 251 | 171 | 31 | 701 | 9 | 239 | 251 | 171 | 31 | 701 |
| Total Number of Endplates | 36 | 956 | 1004 | 684 | 124 | 2804 | 36 | 956 | 1004 | 684 | 124 | 2804 |
| Number of Endplates with Endplate defects | 0 | 32 | 41 | 46 | 24 | 143 | 0 | 106 | 135 | 159 | 39 | 439 |
| Number of Discs with Pfirrmann-1+2 | 11 | 260 | 163 | 68 | 6 | 508 | 16 | 429 | 370 | 134 | 11 | 960 |
| Number of Discs with Pfirrmann-3 | 4 | 134 | 175 | 108 | 10 | 431 | 1 | 34 | 92 | 113 | 15 | 255 |
| Number of Discs with Pfirrmann-4+5 | 3 | 84 | 164 | 166 | 46 | 463 | 1 | 15 | 40 | 95 | 36 | 187 |
| Number of endplates adjacent to disc Pfirrmann-1+2 with Endplate defects | 0 | 1 | 3 | 0 | 0 | 4 | 0 | 74 | 56 | 25 | 1 | 156 |
| Number of endplates adjacent to disc Pfirrmann-3 with Endplate defects | 0 | 4 | 3 | 2 | 0 | 9 | 0 | 16 | 43 | 34 | 5 | 98 |
| Number of endplates adjacent to disc Pfirrmann-4+5 with Endplate defects | 0 | 27 | 35 | 44 | 24 | 130 | 0 | 16 | 36 | 100 | 33 | 185 |

**Table S4a.** The number of discs in the symptomatic group found with central canal stenosis. Results are shown in relation to spinal levels (lower lumbar or upper lumbar), age range and Pfirrmann gradings.

| Symptomatics (OSCLMRIC) |  |  |  |  |  |  |  |  |  |  |  |  |
| --- | --- | --- | --- | --- | --- | --- | --- | --- | --- | --- | --- | --- |
| Age ranges | 30-39 | 40-49 | 50-59 | 60-69 | 70-79 | Total: | 30-39 | 40-49 | 50-59 | 60-69 | 70-79 | Total: |
| Total number of subjects | 121 | 203 | 179 | 155 | 105 | 763 | 121 | 203 | 179 | 155 | 105 | 763 |
| Total number of discs | 242 | 406 | 358 | 310 | 210 | 1526 | 242 | 406 | 358 | 310 | 210 | 1526 |
| Number of discs with Pfirrmann-1,2 | 75 | 115 | 66 | 35 | 9 | 300 | 222 | 349 | 230 | 108 | 17 | 926 |
| Number of discs with Pfirrmann-3 | 64 | 123 | 113 | 89 | 38 | 427 | 15 | 35 | 69 | 90 | 49 | 258 |
| Number of discs with Pfirrmann-4,5 | 103 | 168 | 179 | 186 | 163 | 799 | 5 | 22 | 59 | 112 | 144 | 342 |
| Number of discs with Central Canal Stenosis | 5 | 10 | 24 | 50 | 71 | 160 | 0 | 0 | 6 | 13 | 38 | 57 |
| Number of discs Pfirrmann-1,2 with Central Canal Stenosis | 1 | 0 | 0 | 0 | 1 | 2 | 0 | 0 | 1 | 0 | 0 | 1 |
| Number of discs Pfirrmann-3 with Central Canal Stenosis | 2 | 3 | 5 | 7 | 6 | 23 | 0 | 0 | 0 | 3 | 0 | 3 |
| Number of discs Pfirrmann-4,5 with Central Canal Stenosis | 2 | 7 | 19 | 43 | 64 | 135 | 0 | 0 | 5 | 10 | 38 | 53 |

**Table S4b.** The number of discs in the asymptomatic group found with central canal stenosis. Results are shown in relation to spinal levels (lower lumbar or upper lumbar), age range and Pfirrmann gradings.

| Asymptomatic (TwinsUK) |  |  |  |  |  |  |  |  |  |  |  |  |
| --- | --- | --- | --- | --- | --- | --- | --- | --- | --- | --- | --- | --- |
| MRI Imaging Features | Lower Lumbar (L4/L5, L5/S1) |  |  |  |  |  | Upper Lumbar (L1/2, L2/3) |  |  |  |  |  |
| Age Ranges | 30-39 | 40-49 | 50-59 | 60-69 | 70-79 | Total: | 30-39 | 40-49 | 50-59 | 60-69 | 70-79 | Total: |
| Total Number of Subjects | 9 | 239 | 251 | 171 | 31 | 701 | 9 | 239 | 251 | 171 | 31 | 701 |
| Total Number of Discs | 18 | 478 | 502 | 342 | 62 | 1402 | 18 | 478 | 502 | 342 | 62 | 1402 |
| Number of Discs with central spinal stenosis | 0 | 0 | 6 | 14 | 7 | 27 | 0 | 0 | 6 | 8 | 3 | 17 |
| Number of discs Pfirrmann- 1+2 | 11 | 260 | 163 | 68 | 6 | 508 | 16 | 429 | 370 | 134 | 11 | 960 |
| Number of discs Pfirrmann- 3 | 4 | 134 | 175 | 108 | 10 | 431 | 1 | 34 | 92 | 113 | 15 | 255 |
| Number of discs Pfirrmann- 4+5 | 3 | 84 | 164 | 166 | 46 | 463 | 1 | 15 | 40 | 95 | 36 | 187 |
| Number of discs Pfirrmann- 1+2 with central spinal stenosis | 0 | 0 | 0 | 0 | 0 | 0 | 0 | 0 | 0 | 0 | 0 | 0 |
| Number of discs Pfirrmann- 3 with central spinal stenosis | 0 | 0 | 2 | 6 | 1 | 9 | 0 | 0 | 0 | 1 | 0 | 1 |
| Number of discs Pfirrmann- 4+ with central spinal stenosis | 0 | 0 | 4 | 8 | 6 | 18 | 0 | 0 | 2 | 7 | 3 | 12 |

**Table S5:**
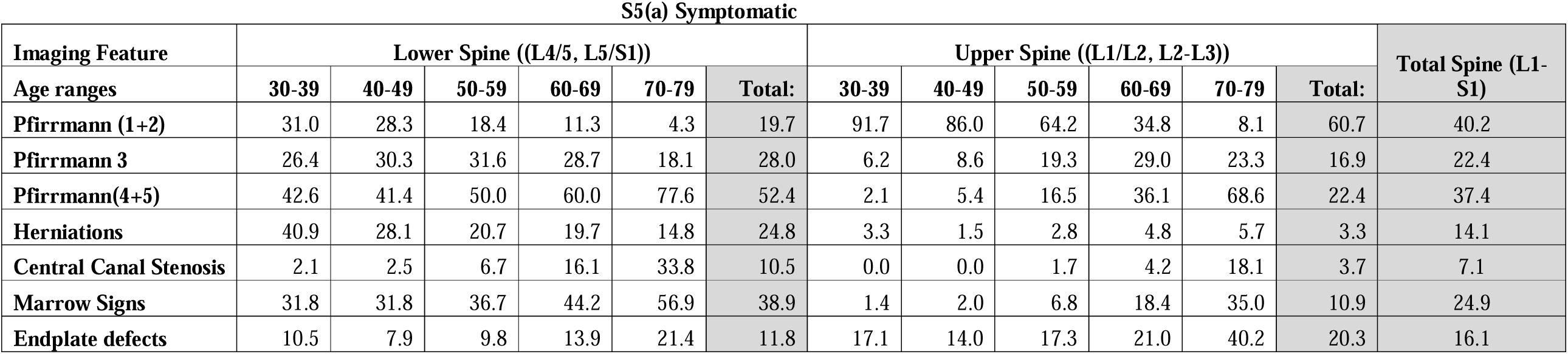
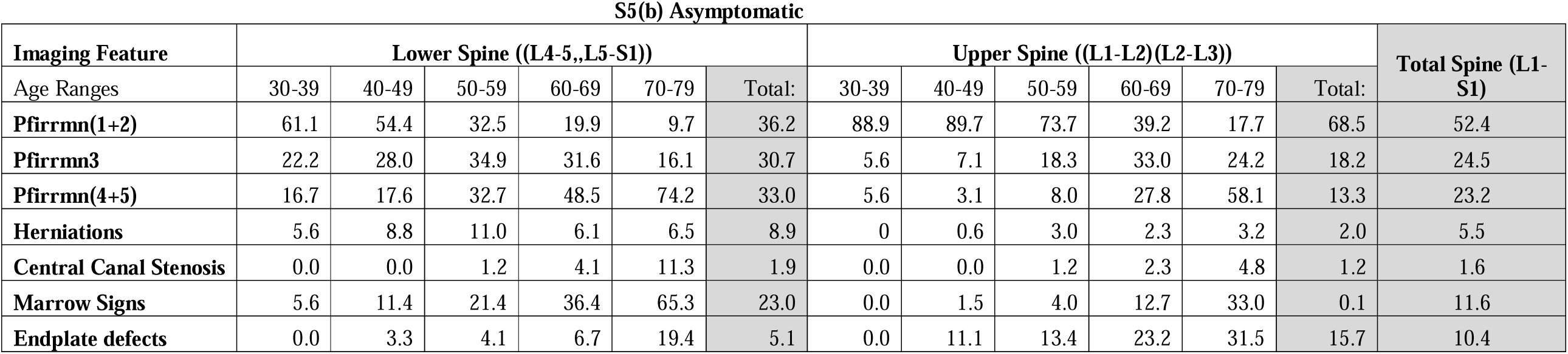
Prevalences of degenerative features in regard to age and spinal level for symptomatic (a) and asymptomatic (b) subjects.

**Table S6.**
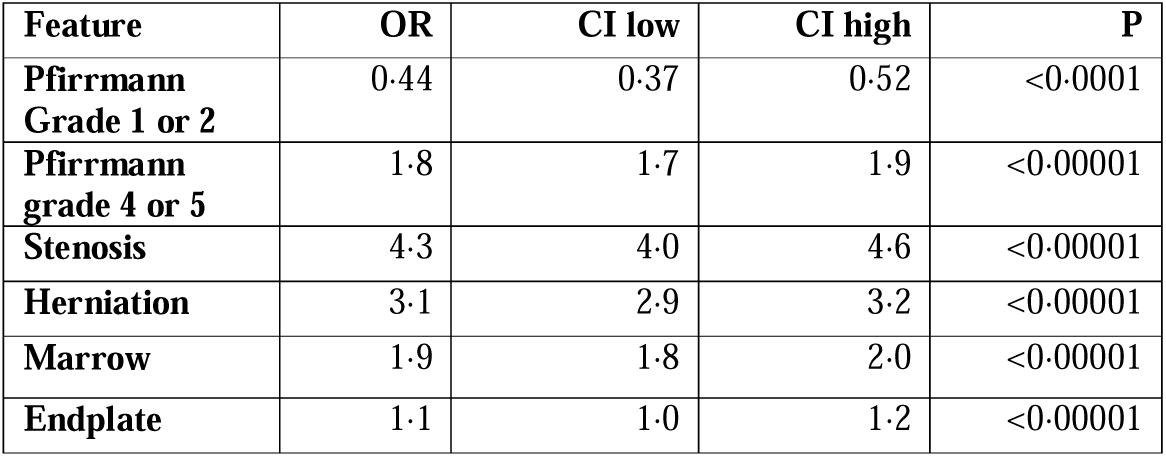
Odds ratios (OR) (symptomatic/asymptomatic), confidence intervals (CI) and probability estimates (P), values for each feature, without taking into account age or spinal level.

**S6b(i).** Disc degeneration Pfirrmann grade 1 or 2 (normal)

| Age band decade (years) | OR | CI low | CI high | P |
| --- | --- | --- | --- | --- |
| 30-39 | 0.3 | 0.1 | 0.8 | =0.01 |
| 40-49 | 0.3 | 0.3 | 0.4 | <0.00001 |
| 50-59 | 0.5 | 0.4 | 0.7 | <0.00001 |
| 60-69 | 0.6 | 0.4 | 0.9 | =0.001 |
| 70-79 | 0.4 | 0.1 | 1.1 | =0.07 |

**S6b(ii).**
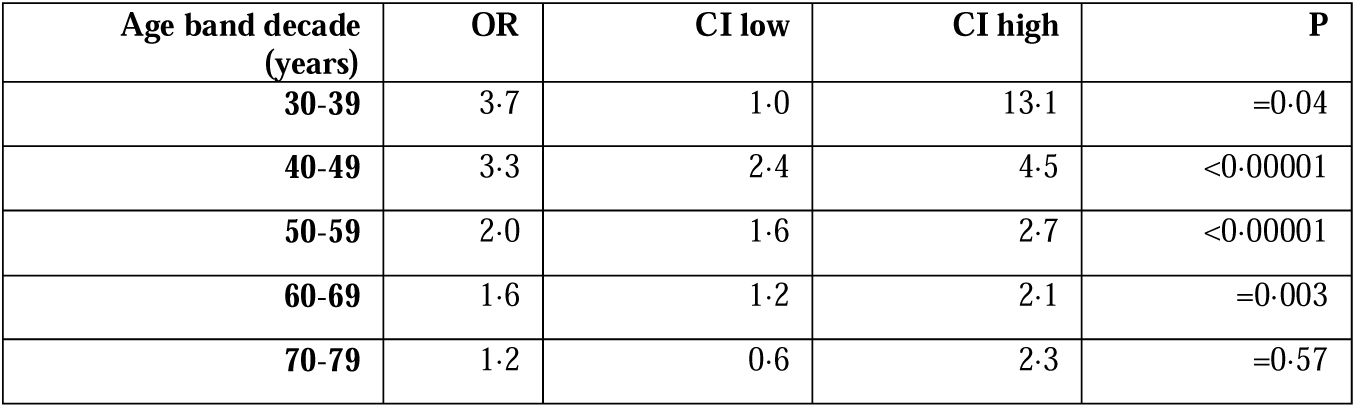
Disc degeneration Pfirrmann grade 4 or 5 (very degenerate):

**S6b(iii).**
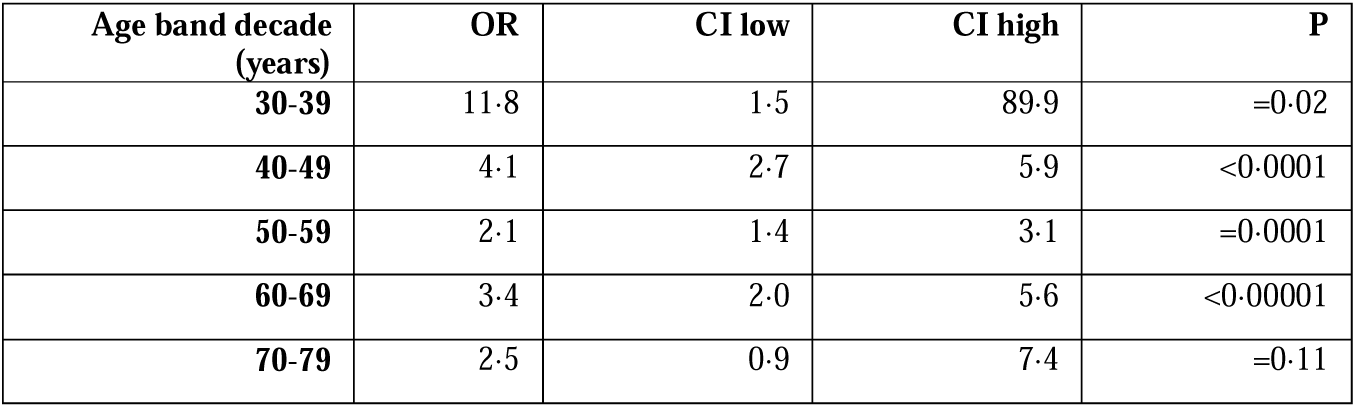
Disc Herniation present.

**S6b(iv).**
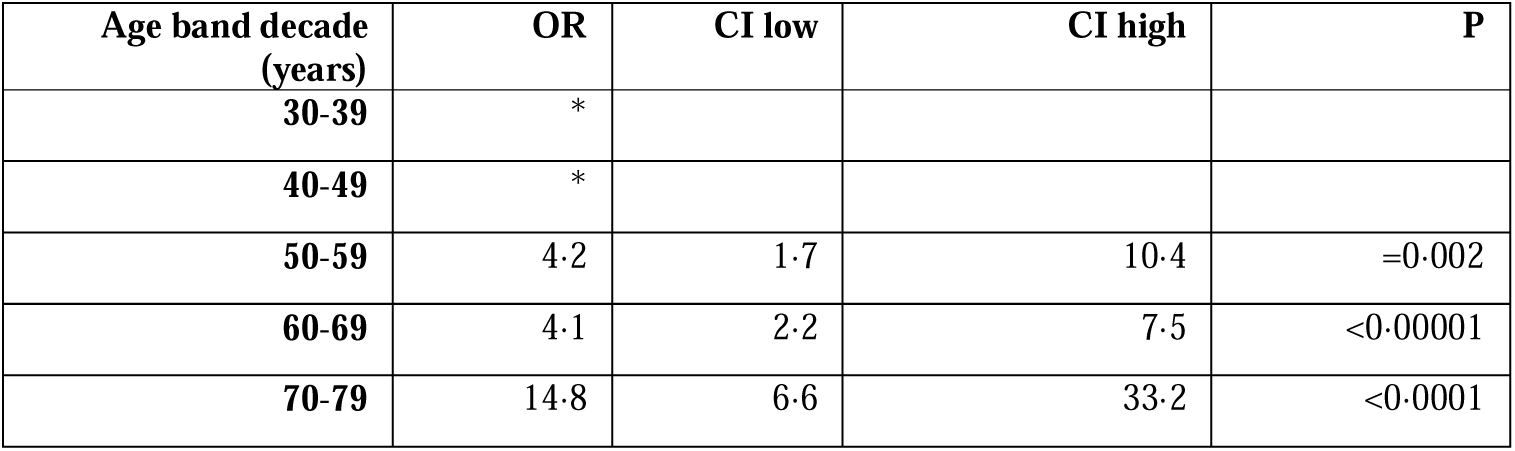
Central Canal stenosis present.

**S6b(v).**
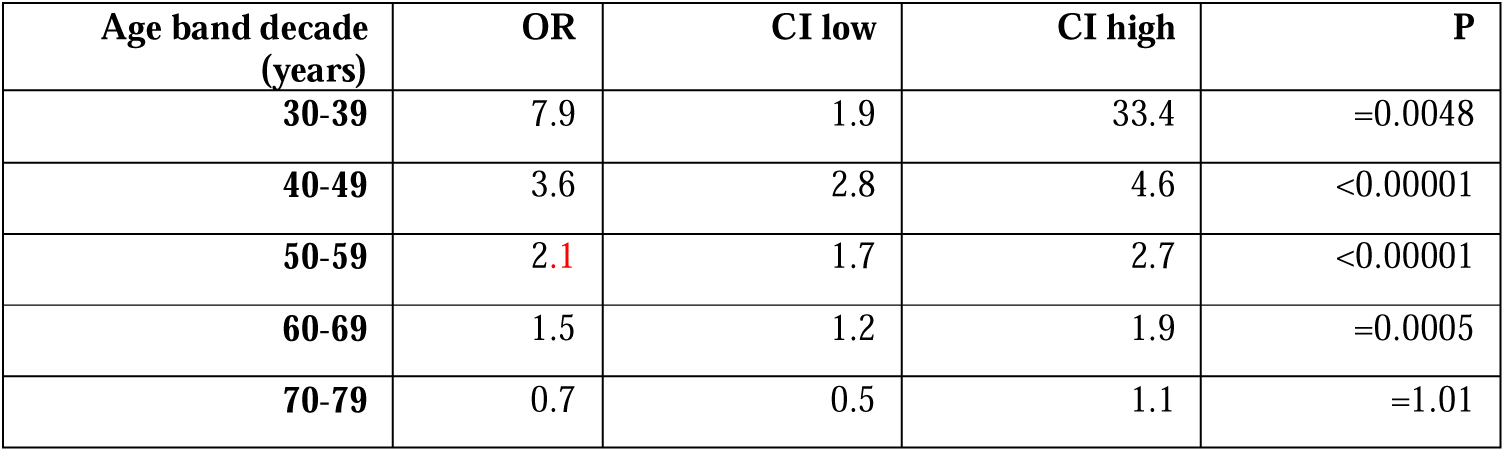
Marrow signs present.

**S6b(vi).**
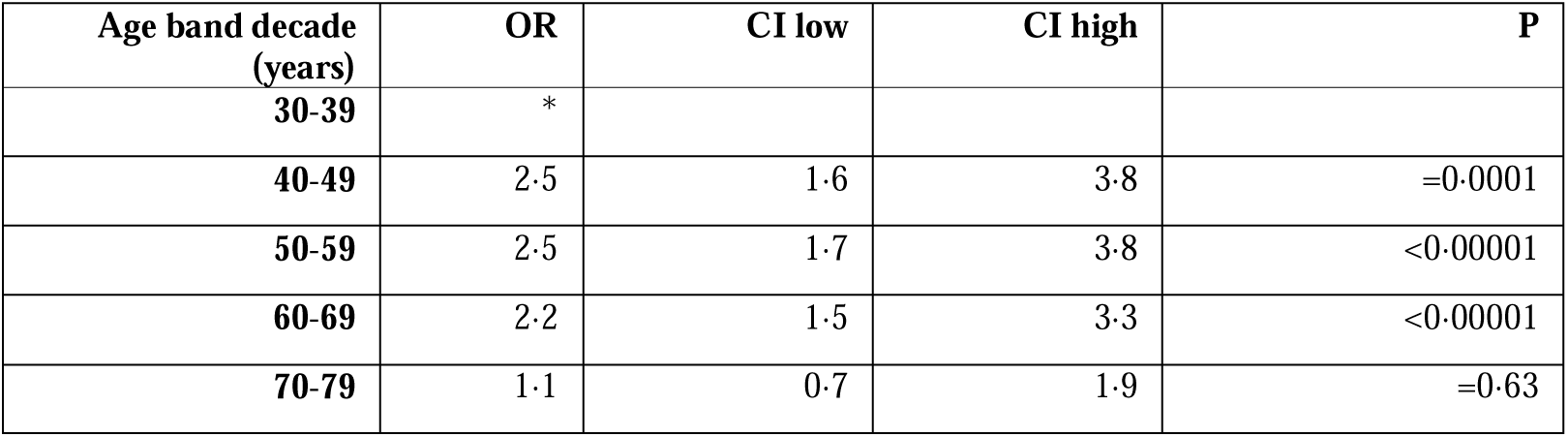
Endplate defects present (Lower lumbar spine)

**S6b(vii).** Endplate defects present (Upper lumbar spine)

S6b(vi) Endplate defects present (Lower lumbar spine)
| Age band decade<br>(years) | OR | CI low | CI high | P |
| --- | --- | --- | --- | --- |
| 30-39 | * |  |  |  |
| 40-49 | 4·2 | 3·2 | 5·7 | <0·00001 |
| 50-59 | 4·6 | 3·5 | 6·2 | <0·00001 |
| 60-69 | 13·6 | 9·3 | 20·0 | <0·00001 |
| 70-79 | 1·5 | 1·0 | 2·2 | 0·05 |

\* - no OR calculated as zero feature positive subjects at that age range in the asymptomatic group

